# Characteristics of Accidental Falls in the Patients with Chronic Kidney Diseases: A 14-Year Retrospective Study and Review

**DOI:** 10.1101/2023.11.29.23299194

**Authors:** Yumei Liao, Li Zhang, Yanmei Peng, Huie Huang, Yuanchang Luo, Jinling Gan, Lina Dong, Yan He, Min Gao, Guang Yang

## Abstract

**Background:** Accidental falls pose a high-risk that should not be overlooked in patients with chronic kidney disease (CKD), as they can result in significant injury or even fatality. This study aimed to investigate the characteristics of CKD patients with fall injuries during hospitalization, discuss potential mechanisms, and to provide an overview of existing prevention methods.

**Methods:** Falls of all patients in our Nephrology ward from 2009 to 2022 were recorded and counted. 48 patients were enrolled. Patient characteristics, injury distribution, cause of fall injury, relevant blood biochemical indicators, and recovery conditions were counted.

**Results:** There were 22,053 hospitalized patients during the study period, with a fall rate of approximately 0.218%. Patients are prone to involuntary falls due to muscle weakness and confusion during nighttime and early morning activities. Injuries are mainly to the head and there is a risk of serious injury and fracture. CKD is associated with anemia, hypertension, water-electrolytes imbalance and secondary hyperparathyroidism. Blood tests showed that patients commonly had anemia, malnutrition, low immunity, as well as abnormal muscle and neuromodulatory ion levels, such as low calcium, low potassium and high phosphorus. Moreover, Patients usually have low blood pressure control ability.

**Conclusion:** Long-term CKD may lead to subjective dysfunction and motor dysfunction by inducing anemia, malnutrition, water-electrolytes imbalance, and blood pressure control ability, thus making patients prone to falls. This study has important implications for hospital ward safety management and fall prevention in CKD patients.

## 1. Introduction

The prevention of accidental falls are critical events that demand attention in the care of patients with long-term CKD, as neglecting them may lead to severe consequences ^1-3^. Chronic kidney disease (CKD) is a kidney disease due to pathological changes or functional reduction of the kidney that lasts for more than 3 months ^4, 5^. Without timely treatment, CKD may gradually progress to end-stage kidney disease (ESKD), which is difficult to treat ^6^. Reportedly, there are approximately 800 million potential patients with CKD worldwide, and this number is probably further increased by global aging and the prevalence of chronic diseases ^7, 8^. Vertigo and weakness are common complications in CKD patients with often suffer sudden falls, severe fractures, and can even lead to death ^9^. Nowadays, advanced nursing care and knowledge sharing are precautery measures for ward safety management; however, it still does not prevent falls occurring. Preventing falls in CKD patients has become an important challenge for ward safety management in the Department of Nephrology.

There are many reasons for the mechanisms associated with CKD-related falls. For example, patients with CKD and ESKD often present with peripheral neuropathy ^10^, which can cause motor dysfunction. The common complications of CKD include anemia, cardiovascular disease, decreased red blood cell survival, iron deficiency, malnutrition, muscle weakness, and osteoporosis due to disturbances in calcium, vitamin D, and phosphate metabolism ^4, 11^. Both anemia and malnutrition can directly contribute to muscle dysfunction ^11, 12^. Anemia and dialysis can lead to cognitive dysfunction by decreasing blood supply to the brain ^13^. These are the reasons why CKD patients are prone to falls. In addition, the fracture rate in CKD patients is 4-5 times higher than that in the healthy population, and the incidence of fractures increases with worsening kidney function ^14, 15^. This could explain the susceptibility of CKD patients to fracture after a fall.

Dialysis is also considered to be one of the reasons for the prone fall in CKD patients. However, this process inevitably results in the loss of small molecule nutrients. Studies have shown that cardiovascular disease, malnutrition, anemia, and abnormal bone metabolism are common complications in patients with maintenance dialysis ^14, 16^. In addition, hemodialysis patients are also prone to muscle cramps and fatigue ^17^. A recent study noted that both hemodialysis and peritoneal dialysis cause decreased muscle mass, increased fat percentage, and depression in patients, however, the symptoms of depression caused by hemodialysis and peritoneal dialysis are different ^18^. Moreover, patients are often accompanied by hypotension or hypertension after hemodialysis treatment ^19, 20^, which may cause symptoms of cerebral ischemia such as dizziness, blurred vision, and syncope. All these factors may lead to sudden falls in patients.

It is often observed in the wards that patients are commonly prone to falls when they get up or walk alone. They stated that the fall was preceded by muscle weakness and confusion, and that even when they realized they could not stand, they did not have the strength to hold on to the support. It seems that almost all the evidence points to subjective dysfunction and motor dysfunction. Therefore, we hypothesize that muscle weakness and sudden dizziness may be the direct cause of the fall, while fracture is the serious consequence after the fall. This study aimed to investigate the characteristics, injury distribution, and high-risk factors in CKD patients who had a fall injury during hospitalization; and to discuss possible mechanisms, and provide an overview of existing prevention methods. For this purpose, we collected information on patients who underwent treatment at our wards from January 2009 to December 2022 and identified 48 patients who had a record of a fall. After that, we counted their history of fall injury, counted etiology, complications, time of fall, degree of injury, and treatment modality. High-risk factor analysis was also performed based on the patients’ personal information and blood biochemical indicators.

## 2. Methods

### 2.1 Ethics

This is a retrospective study and does not address patient harm or additional treatment. The project content and protocol were reviewed and approved by the Ethics Committee of Peking University Shenzhen Hospital (No. 2023-064). All patients signed a pan-informed consent form at the time of hospitalization, and they allowed their consultation data to be used for future research.

### 2.2 Patients

The 48 patients enrolled in the study were the patients who attended the nephrology ward of Peking University Shenzhen Hospital from January 2009 to December 2022. Data on ambulatory patients are not included in the statistics. Inclusion criteria: (i) patients visited Peking University Shenzhen Hospital; (ii) complete visit information was available; (iii) falls during hospital visits. Exclusion criteria: (i) patients had involuntary falls; (ii) patients had other major diseases or genetic disorders affecting muscles and nerves. Rejection criteria: (i) patient had a fall that occurred before the hospital visit.

### 2.3 Surveys

Regular information was collected from patients during their visits. Emphasize the possibility and risk of falls to the patient and family members during the visit. If a fall occurs, treat it promptly. The on-duty nurse was responsible for recording the data, such as time, place, cause, site and extent of injury, and recovery of the patient’s fall.

### 2.4 Blood tests

Patients routinely underwent blood index testing upon admission to the hospital. Venous blood was drawn from the patients and were stored in tubes. Samples for routine blood tests are stored in anticoagulated tubes (#101680720, EDTA-K2-2.0 mg/mL, IMPROVE, China). Blood samples for other assays were stored in procoagulant tubes (separation gel coagulation tube 3.5 mL, IMPROVE, China). Samples were sent to the Department of Laboratory Medicine for uniform testing. The blood routine test was carried out by the Japanese Hisense Macon SYSMEX series blood cell analyzer. Blood biochemical indicators (electrolytes and blood glucose) were detected by Beckmann AU5800 automatic biochemical analyzer. The parathyroid function was detected by Beckmann DXI800 automatic chemiluminescence analyzer. The results of the most recent routine blood test at the time of the patient’s fall will be used.

### 2.5 Blood glucose

Results were based on the patient’s most recent intravenous blood glucose after the fall. Blood glucose was measured by Beckmann AU5800 automatic biochemical analyzer.

### 2.6 Blood pressure

Blood pressure was tested routinely after the patient was admitted to the hospital. Results were based on the blood pressure measured at the time of the patient’s fall.

### 2.7 Statistics

All results are expressed as mean ± standard deviation (SD). The proportion of occurrence is expressed as a percentage. Differences between two groups were calculated using Students’ t-test unpaired tails. GraphPad Prism 6.0 was used for statistics and graphing. *P* < 0.05 represents a significant difference.

## 3. Results

### 3.1 Rate, time, locations, and reasons for falls

To have an overall understanding of fall patients, we counted the rate, time, locations and reasons for falls and performed a correlation analysis (Figure 1). During the study period, a total of 22,053 patients were admitted to our department for inpatient care and 48 falls were recorded. The overall fall rate was less than 0.218%. The time of fall was mainly during 0:00-9:00 Hrs, followed by 12:00-15:00 and 18:00-21:00 Hrs. The locations of falls mainly occurred in wards, hospital corridors and bathrooms, and a few patients fell outside the hospital. The main reasons for falls were getting up, followed by walking, bathroom activities, and turning over while sleeping in bed. Since more patients fell at night (22:00-6:00 Hrs) and in the morning (6:00-9:00 Hrs), we did a correlation analysis and found that 37.5% of the falls occurred at night when they were unconsciously turning over or moving alone when getting up, and 16.67% of the falls occurred in the morning when they were getting up and moving alone. These results suggest that patients are prone to fall due to postural changes (lying, squatting, sitting, standing) at night or when they move alone.

**Figure 1.**
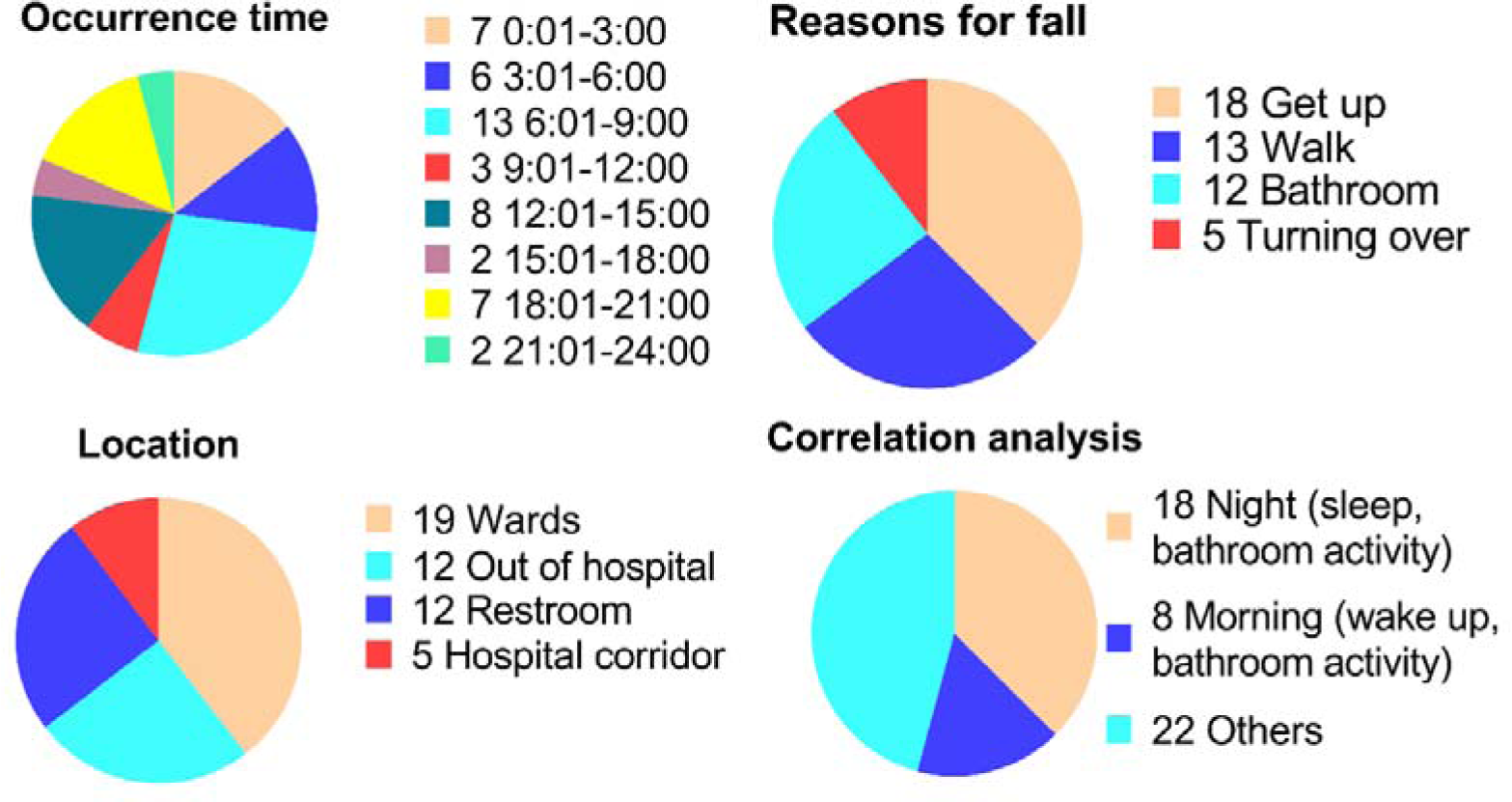
The present study examines the temporal, spatial, and causal factors associated with falls among CKD patients admitted to nephrology wards. A significant majority of falls (more than half) were observed to transpire between 0:00 and 9:00. The primary causes of falls were identified as activities such as getting up, walking, engaging in bathroom-related tasks, and turning over, all of which involve sudden changes in posture. Notably, hospital wards, restrooms, and corridors emerged as the most vulnerable areas in terms of fall occurrences. Furthermore, when considering the correlation among these three factors, it becomes evident that falls are more likely to transpire at night and in the morning when the patients are alone. N = 48, 26 males and 22 females.

### 3.2 The injury site, degree and survival of the falls

To further understand the status of the patients after the fall, we counted the site, degree and survival of the fall injury (Figure 2). Regarding the site of the fall, approximately 18.75% of patients fell but no injuries were observed. In patients with significant injuries, 54.17% had head injuries, 12.5% had hip injuries, 12.5% had hand and arm injuries, and 8.33% had foot and leg injuries. These numbers are more than 100%, mainly because of the presence of multiple tissue injury conditions in the same patient. Further analysis of the degree of injury showed that 37.5% had no obvious trauma, 31.25% had minor abrasions, 18.75% had sprains, swelling, and major trauma, and 10.42% had fractures. Moreover, survival statistics showed that 4 out of 48 patients died. However, of the deceased patients, 2 had no obvious injuries, 1 had minor abrasions, and 1 had a hematoma. These results suggest that patients with CKD are susceptible to head injury after a fall and are still at risk for major trauma or even fracture. In addition, short-term mortality may be independent of the degree of fall injury.

**Figure 2.**
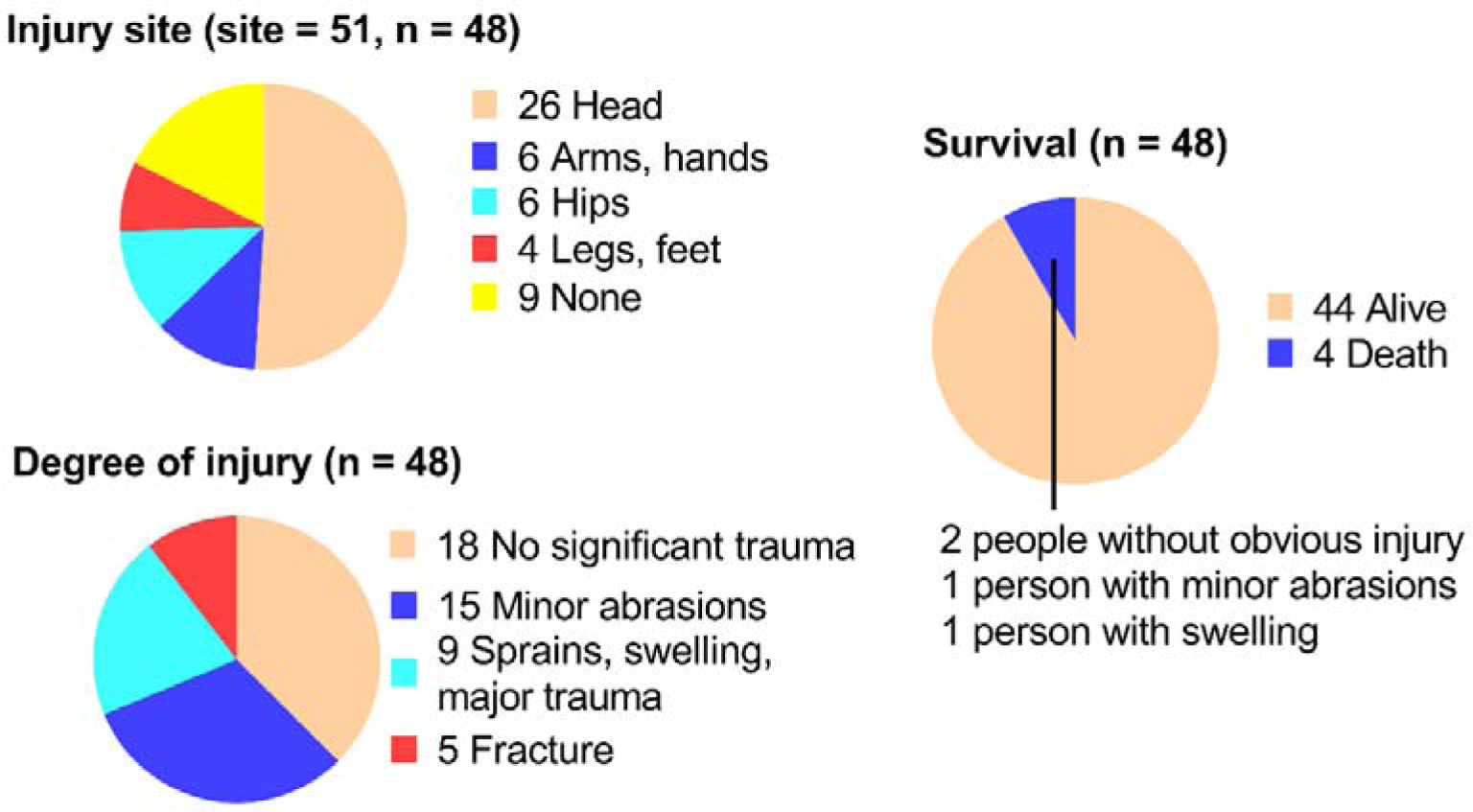
This section focuses on the location, severity, and survival outcomes of fall-related injuries. The head is the most susceptible area to injury in a fall. The majority of cases result in minor or no injuries, while a small proportion exhibit more severe consequences. Approximately one-third of individuals experience more serious injuries or fractures. Mortality rates are below 10%, with half of these cases showing no visible signs of trauma. N = 48, 26 males and 22 females.

### 3.3 Correlation between falls and CKD types, complications, and treatment approaches

To understand the high risk-factors for falls, we counted the primary diseases, complications and treatment approaches (Figure 3). The results showed that a high percentage of patients with falls had nephrotic syndrome, chronic nephritis, and diabetic nephropathy. Among them, 64.58% progressed to CKD stage 5. Moreover, these patients commonly had complications of anemia, water-electrolytes imbalance, hypertension and secondary hyperparathyroidism. Furthermore, some of these patients did not receive dialysis treatment, some received single hemodialysis or peritoneal dialysis, and some received both treatments. These results suggested that patients with combined CKD and hypertension, anemia, secondary hyperparathyroidism, and water-electrolytes imbalance are prone to falls. The effect of treatment approaches on falls could not be determined at this time.

**Figure 3.**
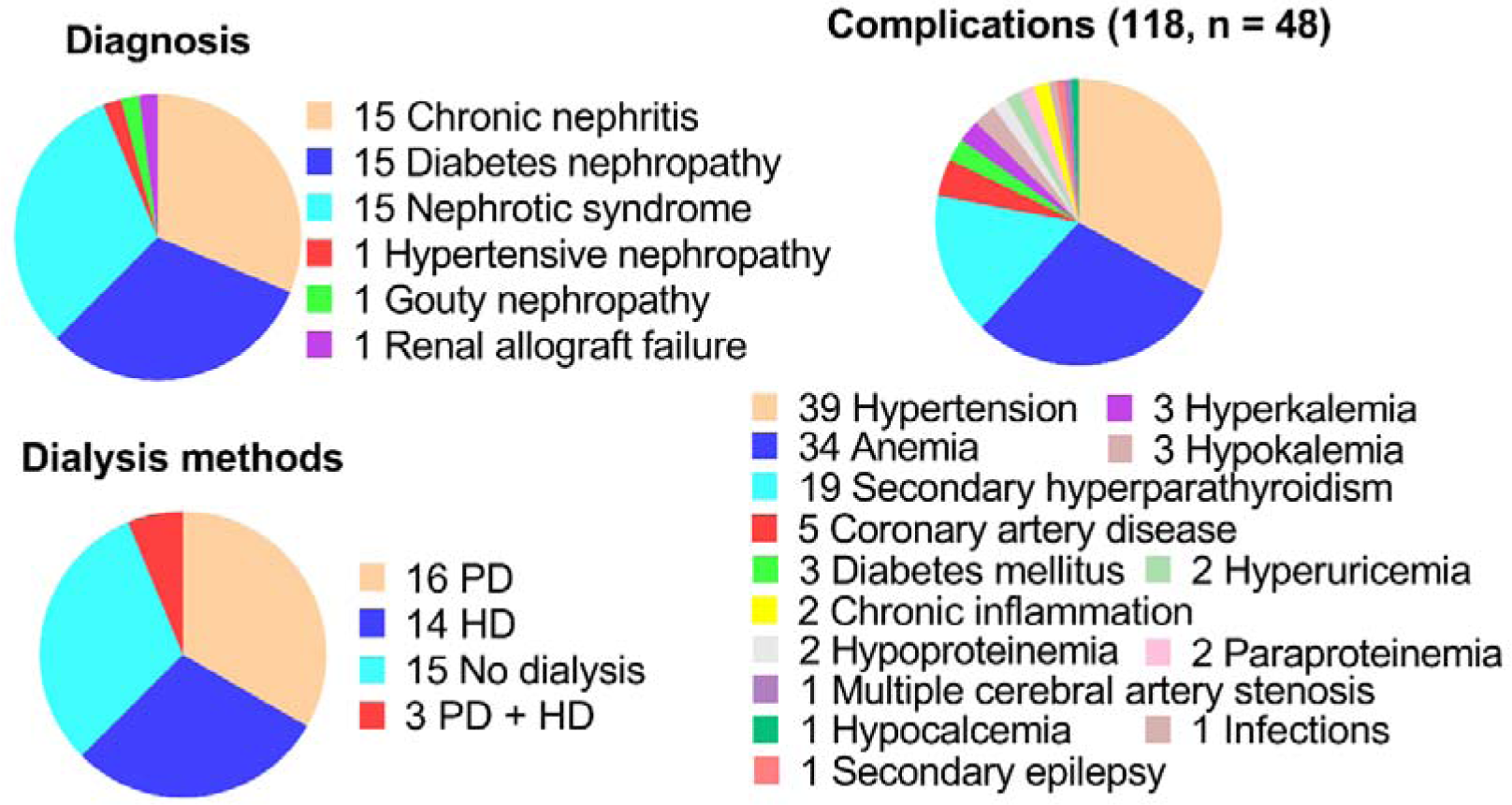
The correlation between falls and primary disease types, complications and treatment approaches. It was found that the high-risk factors were closely related to hypertension, anemia, secondary hyperparathyroidism, and water-electrolytes imbalance. PD, peritoneal dialysis. HD, hemodialysis. N = 48, 26 males and 22 females.

### 3.4 Effects of gender and age on falls

To understand the effects of gender and age on falls, we conducted preliminary statistics (Figure 4). The results showed that the absolute number of falls in men was slightly more than for women. Moreover, the mean age of women was slightly higher than that of men, but there was no statistically significant difference. This may be due to insufficient group size.

**Figure 4.**
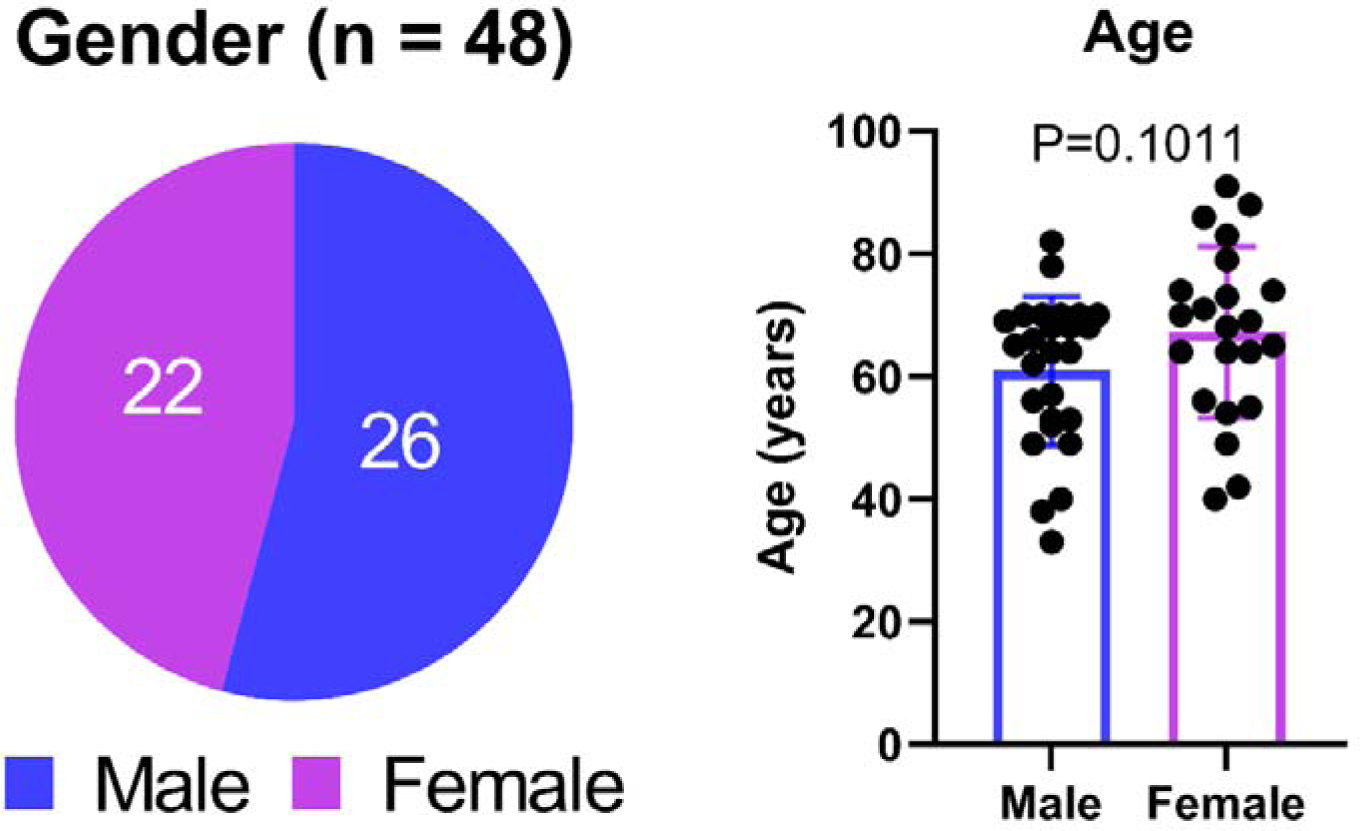
Gender and age statistics of fallen patients. Age and gender did not show significant differences in the effect of falls. (Age, 60.92±12.16 vs. 67.23±13.95, years. N = 48, 26 males and 22 females.)

### 3.5 CKD-related falls are associatedwith malnutrition and blood homeostasis imbalance

The above results showed that the patients commonly experienced sudden weakness and fainting during the transition between lying, sitting, and standing positions followed by involuntary falls. We hypothesized that it was likely to be associated with subjective dysfunction and motor dysfunction, and subsequently we measured the relevant factors levels in the blood. The results of blood indicators are presented separately for men and women (Figure 5). NRV is the normal reference value range. The results showed no significant differences between men and women in all indicators. However, if compared with NRV, patients generally had low hemoglobin, low blood albumin, high blood calcium, high blood phosphorus, and high parathormone; some patients had low blood potassium; a few patients had low blood sodium, and high blood glucose.

**Figure 5.**
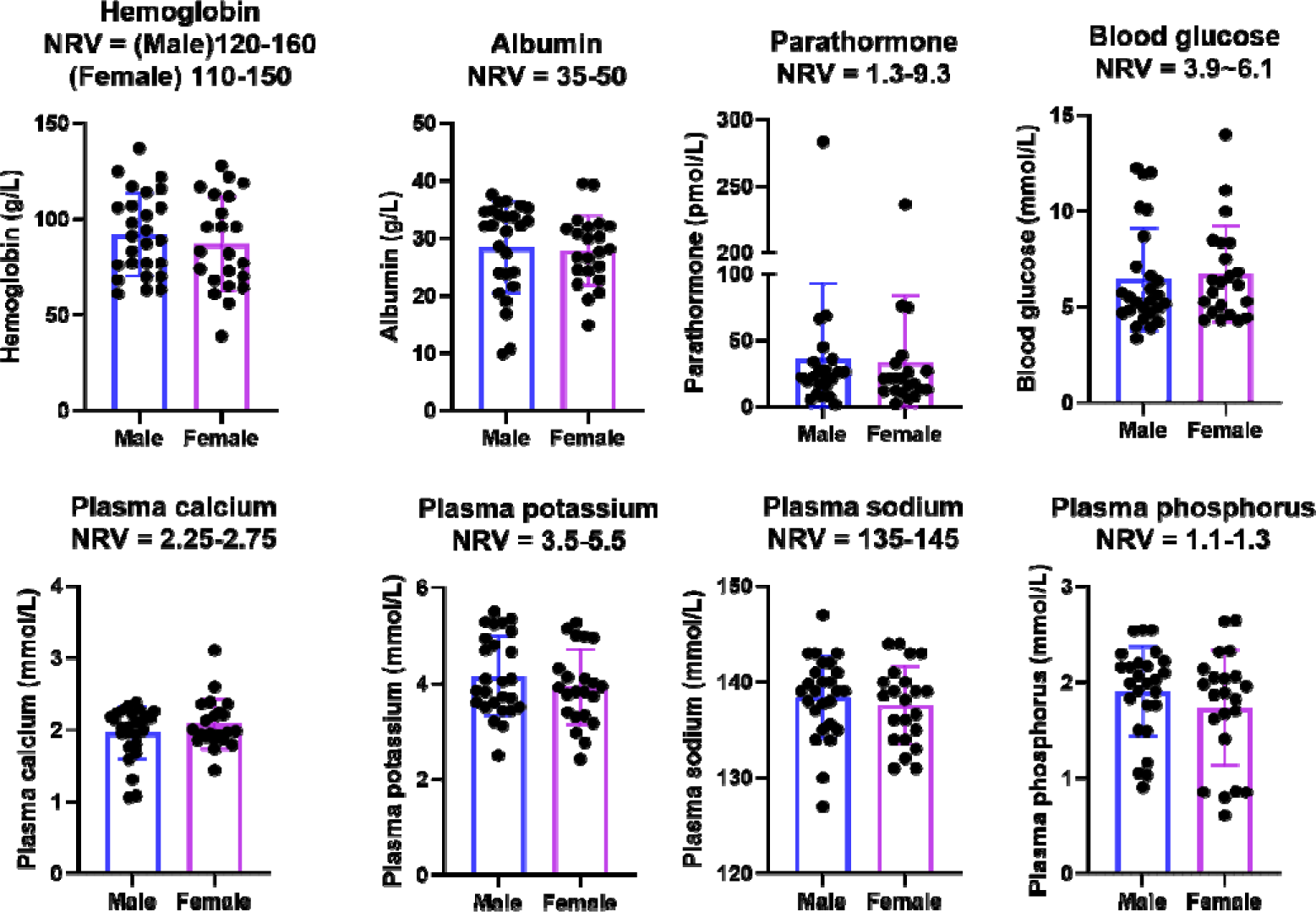
Patient’s blood biochemical indicators (hemoglobin, albumin, potassium, phosphorus, calcium, parathormone,sodium, and blood glucose). No significant differences were observed between men and women in all the indicators examined. (Hemoglobin, 92.15±21.62 vs. 87±24.63, g/L. Albumin, 28.42±7.995 vs.27.87±6.061, g/L. Parathormone, 36.46±56.44 vs. 33.63±50.31, pmol/L. Blood glucose, 6.448±2.679 vs. 6.734±2.502, mmol/L. Plasma calcium, 1.964±0.3627 vs. 2.08±0.3469, mmol/L. Plasma potassium, 4.155±0.8321 vs. 3.921±0.7901, mmol/L. Plasma sodium, 138.4±4.286 vs. 137.6±4.076, mmol/L. Plasma phosphorus, 1.909±0.4686 vs. 1.733±0.6004, mmol/L. N = 48, 26 males and 22 females.) However, it was noted that patients generally exhibited low levels of hemoglobin and blood albumin, alongside elevated levels of blood calcium, blood phosphorus, and parathormone. Some patients also presented with low blood potassium, while a few showed low blood sodium and high blood glucose. NRV, normal reference value.

### 3.6 CKD-related falls are associated with low blood pressure control ability

Abnormal blood pressure and low blood pressure control ability (LBPCA) can cause dizziness. We analyzed systolic blood pressure, diastolic blood pressure, and pulse pressure (Figure 6). The results showed that the patients generally had high blood pressure and a large difference in pulse pressure, which means LBPCA.

**Figure 6.**
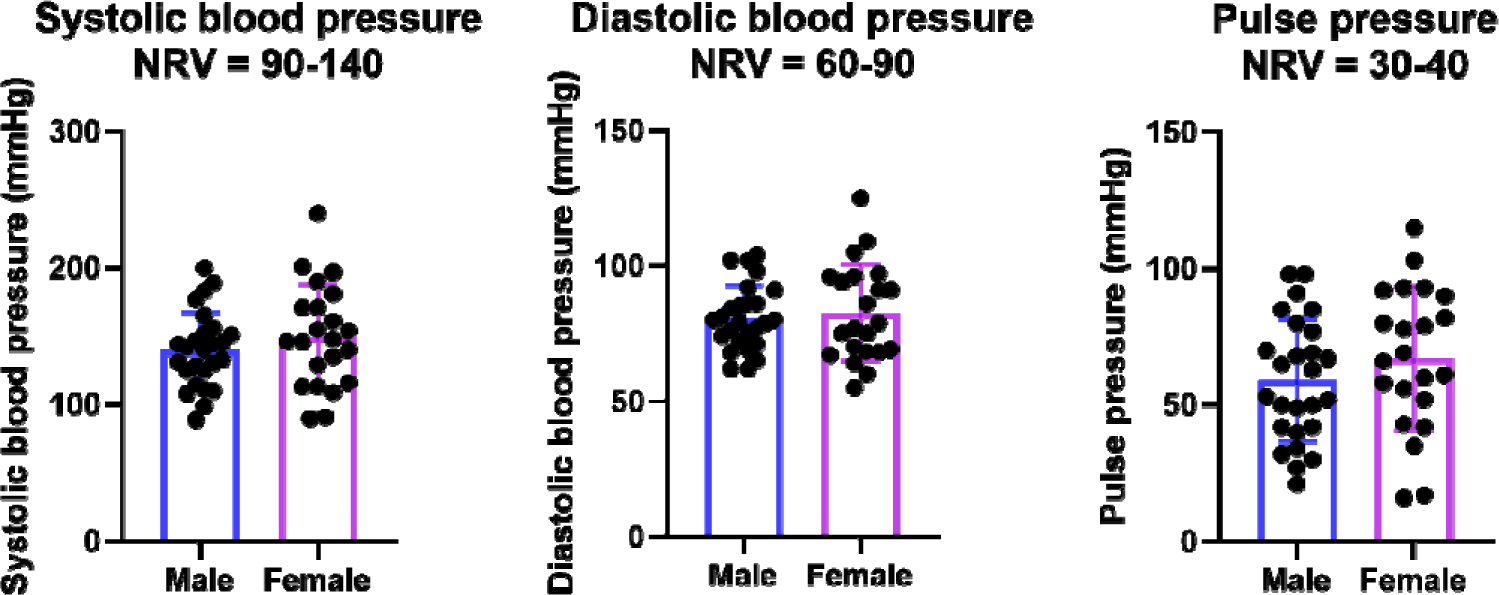
Patients’ systolic blood pressure, diastolic blood pressure, and pulse pressure. CKD patients generally had high blood pressure and a large difference in pulse pressure NRV, normal reference value. (Systolic blood pressure, 139.9±27.58 vs. 149.9±37.78, mmHg. Diastolic blood pressure, 80.77±12 vs. 82.59±17.78, mmHg. Pulse pressure, 59.15±22.4 vs. 67.27±26.37, mmHg. N = 48, 26 males and 22 females.)

## 4. Discussion

In recent decades, it has been gradually realized that CKD patients are prone to falls and fractures. To prevent patients from having a lower quality of life and a higher financial burden, many ways have been proposed to improve ward safety management, such as enhanced nursing care and knowledge promotion. This has reduced the occurrence of falls to some extent, but not completely prevent them. In this study, we conducted a systematic study and found that patients are prone to falls at night and morning during postural changes due to muscle weakness and confusion (Figure 7). CKD is associated with anemia, water-electrolytes disturbances, hypertension, and secondary hyperparathyroidism. These fallen patients also commonly have anemia, malnutrition, LBPCA, and abnormal muscle- and neuro-modulatory ion levels. This study hopes to raise awareness of the potential for fall-related safety emergencies to occur despite existing safety management conditions in nephrology wards and to take relevant precautions.

**Figure 7.**
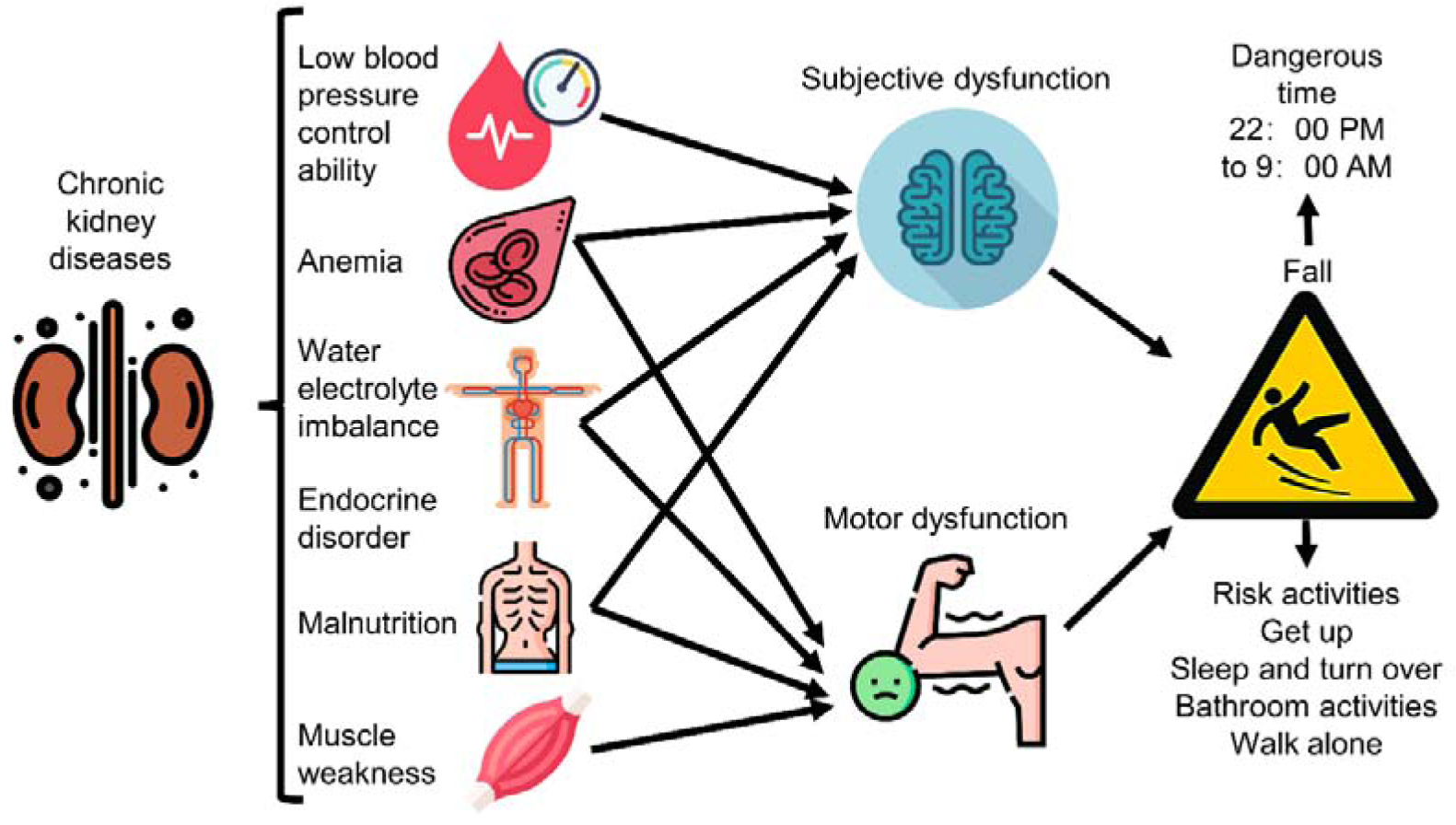
Mechanisms and risk analysis of fall-prone patients with chronic kidney disease. Copyright explanation: Icons are free files from Flaticon website and WPS office, personal use is allowed for free. The final images are made by us. Please refer to the Acknowledgments section for more details.

### 4.1 Reasons for fall-prone patients with CKD

There are many types of CKD, and although this study found a larger proportion of absolute number of falls in patients with nephrotic syndrome, chronic nephritis, and diabetic nephropathy. The base of such CKD patients is large and the proportion of falls in all patients has not been calculated. Therefore, we were unable to determine the relationship between the type of CKD and falls. However, all of these diseases are associated with osteoporosis, muscle weakness, and subjective dysfunction. For example, type 2 diabetes causes reduced cognitive function, muscle strength, and level of performance capacity ^21-23^. Studies based on childhood nephrotic syndrome have shown that nephrotic syndrome predisposes children to malnutrition, putting them at risk for growth retardation, muscle wasting, and cognitive impairment ^24, 25^. Similarly, clinical adult studies have shown that ESKD patients on hemodialysis have significantly less nutritional status and muscle mass than healthy groups ^26^. These studies illustrate that all CKD-related diseases may lead to malnutrition, subjective dysfunction, and motor dysfunction.

Subsequent results showed that anemia, hypertension, secondary hyperparathyroidism, and water-electrolytes disturbances were the most prevalent complications among fall patients. Anemia causes muscles and nerves dysfunction which may lead to falls in patients ^12, 13^. The presence of anemia was also confirmed by the low hemoglobin of the patients in this study. Parathyroid hormone disorders are associated with increased risk of muscle dysfunction and fractures; and hyperparathyroidism is also a high-risk factor among CKD ^27^. Here, patients were found to have hyperparathyroidism at the time of diagnosis, which was further confirmed in subsequent blood tests.

In addition, rapid changes in blood pressure is also strongly associated with subjective dysfunction ^28^. High blood pressure is common in patients with CKD, while the blood pressure usually decreases after dialysis, with a few cases of elevated blood pressure. It is known that both hypertension and hypotension may cause dizziness. In the present study, patients generally had high blood pressure and LBPCA. Considering the weakened cardiovascular regulation in the elderly CKD group, such frequent fluctuations in blood pressure may be more harmful to them. Therefore, prolonged large fluctuations and orthostatic hypotension in blood pressure may lead to sudden dizziness.

Moreover, blood biochemical parameters are closely related to health. Here, patients generally had low blood calcium, high blood phosphorus, and in some cases low blood potassium. Abnormalities in these ion concentrations can lead to abnormal muscle and nerve function. For example, calcium ions are involved in muscle contraction, muscle construction, and nerve signaling ^29^. High blood phosphorus increases inflammation, anemia, and skeletal muscle atrophy ^30^. Additionally, although the patient’s blood potassium is within the normal range, however, it is still considered the low concentration (<4 mmol/L). This is also detrimental to the maintenance of normal function of nerve and muscle cells and cardiac muscle cells.

Furthermore, CKD is closely associated with age, and aging leads to functional decline through increased kidney susceptibility ^31^. Aging causes not only a decline in kidney function but also a decline in systemic physiological functions, such as muscle atrophy, osteoporosis, and subjective impairment of consciousness ^32^. This makes it understandable why older CKD groups are more prone to falls and fractures. Consistent with this view, fallen patients are predominantly in the age above 50 years of age group.

### 4.2 Fall prevention strategies

Nowadays, there are many measures to prevent falls in CKD patients, which mainly include assessment and safety management. Assessment refers to asking patients about their basic conditions, checking all physiological indicators as well as understanding dialysis-related conditions. After that, the high risk-factors for falls are assessed and studied. Safety management is the training of nursing staff including organizing discussions, developing a fall prevention process, and taking effective preventive measures. Moreover, regular emergency plan training drills and knowledge dissemination to patients, family members, and companions are conducted. These approaches enable everyone to focus on fall prevention and bring down the incidence of falls.

The guidlines implement are: (i) Fall risk assessment for each patient upon admission to the hospital and every Monday and Thursday. (ii) Carefully explain the risks and dangers of falls and preventive measures. (iii) Create a fall prevention video to be played on a loop in the lobby of the ward. (iv) Make a colorful picture book with illustrations for patients and their families to read. (v) Periodically, a grip strength meter is used to assess the patient’s muscle strength. It is worthwhile to be alerted when the value is too low. These approaches are simple and effective. If a fall occurs, the entire staff needs to discuss the cause of the occurrence and propose targeted preventive measures. These measures have greatly reduced the occurrence of falls in our wards and have kept the rate of falls among inpatients in our department at a low level.

However, the present study still found some deficiencies in the existing prevention strategies. First, it is found that patients are prone to fall out of bed during nighttime sleep and turn over. This problem could be effectively addressed if bedside guardrails could be raised. Even if the patient is homebound, it is recommended to use bed with guardrails. Never put down the bed rails on the bed of the patient himself to prevent the patient’s body from being brought down by the gravitational inertia of the bed rails resulting in a fall. Second, the patient’s activity alone at night is also an important reason for falls. Through conversation with the patient, we learned that the patient did not want to disturb others at night and preferred to go to the bathroom by himself. The reasons for such falls can be attributed to occurring in the absence of a chaperone and a sudden increase in movement. Therefore, it isrecommended that patients must be equipped with a walking aid or exoskeleton as a better solution. In addition, patients should avoid violent postural changes. Third, patients are also prone to fall when getting up, which is directly related to postural changes. It takes time for the body to adapt to the new physiological state from waking up to standing up. Older CKD patients are already weak and their bodies cannot withstand the rapid change in physiological status. Fourth, patients are prone to fall when walking outside. For such patients, exoskeletons and walking aids are the best choices.

Many scholars have suggested that early intervention therapy is an effective strategy to prevent falls and fractures. Covino et al. suggested that intervention therapy to prevent osteoporosis is an effective way to reduce falls and fractures in CKD patients ^15^. Shaker et al. found that when hemoglobin recovered to 115-125 g/L, CKD patients obtained significant improvement in brain blood supply and cognitive function. Stroke, cardiovascular disease and hypertensive complications disappeared when hemoglobin exceeded 125 g/L. Nutritional interventions to improve malnutrition in CKD patients are also considered a practical and effective way to significantly improve muscle strength ^26, 33^. In addition, physical interventions have been shown to be effective. Kesik et al. demonstrated that the use of both hot and cold compresses on the extremities reduced both comfort during treatment and post-treatment muscle spasm and fatigue in hemodialysis patients, and that hot compresses were significantly more effective than cold compresses ^17^.

Recently, emerging biological therapies, such as stem cells and exosomeshave been recognized as effective approaches to promote muscle regeneration, bone regeneration, kidney diseases, and neurological repair. Moreover, engineering strategies enable targeted modification of cells and exosomes so that they can be more functional at the same time ^34^. However, there are no studies on CKD combined with subjective dysfunction and muscle dysfunction. These biological therapies deserve further exploration.

### 4.3 Limitations

There are some limitations in this study. First, the number of cases is limited, and multi-center studies are needed to expand data sources in the future. Second, there is a lack of proportional analysis of patients who fall and all CKD patients. Third, the effect of dialysis modalities on falls is uncertain. These issues are mainly limited by the group size. Fourth, the blood data is still limited, and we plan to find more mechanisms through high-throughput sequencing and multi-omics in the future. Fifth, there is a lack of intervention trials to find prevention options to reduce falls. Sixth, we observed that deaths after falls were not proportional to the degree of fall injuries, and the reason is unclear.

## 5. Conclusions

In conclusion, long-term CKD may lead to sudden vertigo and muscle dysfunction through anemia, malnutrition, water-electrolytes imbalance, and LBPCA, thereby increasing the chance of falls and fractures (Figure 7). Additionally, avoiding patients acting alone, paying attention to night care, and telling patients to avoid sudden changes in posture are key to avoiding falls. This study has important implications for ward safety management and prevention of falls in CKD patients. In the future, we will find ways to reduce the chance of falling through preclinical and clinical studies to find nutritional interventions.

## Data Availability

All data produced in the present study are available upon reasonable request to the authors.

## 6. Abbreviations

CKD: Chronic kidney disease
ESKD: end-stage kidney disease
NRV: normal reference value
LBPCA: low blood pressure control ability

## 7. Acknowledgements

The authors express their gratitude to all colleagues in clinical departments and laboratories for their valuable contributions to this study. Additionally, we extend our sincere appreciation to the website Flaticon. We would like to extend special thanks to the talented iconographers Freepik, Eucalyp, Juice Fish, Flatart_icons, and Flat_icons for their provision of a wide array of visually appealing icons.

## 9. Contribution

Conceptualization: YL and GY. Methodology: YL, LZ, YP, HH, YLuo, JG, LD, YH, and MG. Validation: YL, LZ, YP, HH, YLuo, JG, LD, YH, and MG. Formal analysis: YL and GY. Investigation: YL, LZ, YP, HH, YLuo, JG, LD, YH, and MG. Data curation: YL and GY. Supervision: ZuX, WL and ZiX. Funding acquisition: WL, ZiX, GY, and ZuX. All authors contributed to the writing. All authors have read and agreed to the published version of the manuscript.

## 11. Data availability

The relevant raw data are provided in the annex.

## 12. References

1 Kimura A, Paredes W, Pai R, Farooq H, Buttar RS, Custodio M, et al. Step length and fall risk in adults with chronic kidney disease: a pilot study. BMC Nephrol. 2022;23(1): 74.

2 Heybeli C, Kazancioglu R, Smith L, Veronese N, Soysal P. Risk factors for high fall risk in elderly patients with chronic kidney disease. Int Urol Nephrol. 2022;54(2): 349–356.

3 Kistler BM, Khubchandani J, Jakubowicz G, Wilund K, Sosnoff J. Falls and Fall-Related Injuries Among US Adults Aged 65 or Older With Chronic Kidney Disease. Prev Chronic Dis. 2018;15: E82.

4 Webster AC, Nagler EV, Morton RL, Masson P. Chronic Kidney Disease. Lancet. 2017;389(10075): 1238–1252.

5 Yang G, Tan L, Yao H, Xiong Z, Wu J, Huang X. Long-Term Effects of Severe Burns on the Kidneys: Research Advances and Potential Therapeutic Approaches. Journal of Inflammation Research. 2023;16: 1905–1921.

6 Zhao Z, Qiao H, Ge Y, Kannapel CC, Sung SJ, Gaskin F, et al. Autoimmune experimental orchitis and chronic glomerulonephritis with end stage renal disease are controlled by Cgnz1 for susceptibility to end organ damage. Clin Immunol. 2021;224: 108675.

7 Kovesdy CP. Epidemiology of chronic kidney disease: an update 2022. Kidney Int Suppl (2011). 2022;12(1): 7-11.

8 Hill NR, Fatoba ST, Oke JL, Hirst JA, O’Callaghan CA, Lasserson DS, Hobbs FD. Global Prevalence of Chronic Kidney Disease - A Systematic Review and Meta-Analysis. PLoS One. 2016;11(7): e0158765.

9 Sarad N, Jannath SY, Ogami T, Khedr S, Omar H, Thorson T, Kopp M. Chronic kidney disease and polypharmacy as risk factors for recurrent falls in a nursing home population. Health Sci Rep. 2023;6(10): e1564.

10 Li X, Sun H, Zhang Z, Liu J, Xu H, Ma L, et al. Shear Wave Elastography in the Diagnosis of Peripheral Neuropathy in Patients With Chronic Kidney Disease Stage 5. Front Endocrinol (Lausanne). 2022;13: 899822.

11 Wang K, Liu Q, Tang M, Qi G, Qiu C, Huang Y, et al. Chronic kidney disease-induced muscle atrophy: Molecular mechanisms and promising therapies. Biochem Pharmacol. 2023;208: 115407.

12 Gonçalves CEA, Silva PO, Soares MS, Bunn PS, Lima CMA, Lopes AJ. Muscle dysfunction is associated with poorer health-related quality of life in adults with sickle cell anaemia. J Back Musculoskelet Rehabil. 2019;32(1): 43–53.

13 Shaker AM, Mohamed OM, Mohamed MF, El-Khashaba SO. Impact of correction of anemia in end-stage renal disease patients on cerebral circulation and cognitive functions. Saudi J Kidney Dis Transpl. 2018;29(6): 1333–1341.

14 Abdalbary M, Sobh M, Elnagar S, Elhadedy MA, Elshabrawy N, Abdelsalam M, et al. Management of osteoporosis in patients with chronic kidney disease. Osteoporos Int. 2022;33(11): 2259–2274.

15 Covino M, Vitiello R, De Matteis G, Bonadia N, Piccioni A, Carbone L, et al. Hip Fracture Risk in Elderly With Non-End-Stage Chronic Kidney Disease: A Fall Related Analysis. Am J Med Sci. 2022;363(1): 48–54.

16. Murdeshwar HN, Anjum F. Hemodialysis. StatPearls. Treasure Island (FL): StatPearls Publishing Copyright © 2022, StatPearls Publishing LLC., 2022.

17 Kesik G, Ozdemir L, Yıldırım T, Jabrayilov J, Çeliksöz G. Effects of warm or cold compresses applied to the legs during hemodialysis on cramps, fatigue, and patient comfort: A placebo-controlled randomized trial. Hemodial Int. 2023.

18 Vučković M, Radić J, Kolak E, Nenadić DB, Begović M, Radić M. Body Composition Parameters Correlate to Depression Symptom Levels in Patients Treated with Hemodialysis and Peritoneal Dialysis. Int J Environ Res Public Health. 2023;20(3).

19 Zoccali C, Tripepi G, Neri L, Savoia M, Baro’ Salvador ME, Ponce P, et al. Effectiveness of Cold Hemodialysis (HD) for the Prevention of HD Hypotension and Mortality in the General HD Population. Nephrol Dial Transplant. 2023.

20 Van Buren PN, Inrig JK. Hypertension and hemodialysis: pathophysiology and outcomes in adult and pediatric populations. Pediatr Nephrol. 2012;27(3): 339–350.

21 Midorikawa M, Suzuki H, Suzuki Y, Yamauchi K, Sato H, Nemoto K, et al. Relationships between Cognitive Function and Odor Identification, Balance Capability, and Muscle Strength in Middle-Aged Persons with and without Type 2 Diabetes. J Diabetes Res. 2021;2021: 9961612.

22 Travis C, Srivastava PS, Hawke TJ, Kalaitzoglou E. Diabetic Bone Disease and Diabetic Myopathy: Manifestations of the Impaired Muscle-Bone Unit in Type 1 Diabetes. J Diabetes Res. 2022;2022: 2650342.

23 Zhang PN, Zhou MQ, Guo J, Zheng HJ, Tang J, Zhang C, et al. Mitochondrial Dysfunction and Diabetic Nephropathy: Nontraditional Therapeutic Opportunities. J Diabetes Res. 2021;2021: 1010268.

24 Gehad MH, Yousif YM, Metwally MI, AbdAllah AM, Elhawy LL, El-Shal AS, Abdellatif GM. Utility of muscle ultrasound in nutritional assessment of children with nephrotic syndrome. Pediatr Nephrol. 2022.

25 Solarin A, Adekunle MO, Olutekunbi O, Lamina O, Njokanma F. Nutritional Assessment of Children with Nephrotic Syndrome in a tertiary institution: A case controlled study. 2019.

26 Sabatino A, Regolisti G, Delsante M, Di Motta T, Cantarelli C, Pioli S, et al. Noninvasive evaluation of muscle mass by ultrasonography of quadriceps femoris muscle in End-Stage Renal Disease patients on hemodialysis. Clin Nutr. 2019;38(3): 1232–1239.

27 Romagnoli C, Brandi ML. Muscle Physiopathology in Parathyroid Hormone Disorders. Front Med (Lausanne). 2021;8: 764346.

28 Rossi DM, de Souza HCD, Bevilaqua-Grossi D, Vendramim ACC, Philbois SV, Carvalho GF, et al. Impairment on Cardiovascular Autonomic Modulation in Women with Migraine. Int J Environ Res Public Health. 2022;20(1).

29 Berchtold MW, Brinkmeier H, Müntener M. Calcium ion in skeletal muscle: its crucial role for muscle function, plasticity, and disease. Physiol Rev. 2000;80(3): 1215–1265.

30 Czaya B, Heitman K, Campos I, Yanucil C, Kentrup D, Westbrook D, et al. Hyperphosphatemia increases inflammation to exacerbate anemia and skeletal muscle wasting independently of FGF23-FGFR4 signaling. Elife. 2022;11.

31 Sato Y, Yanagita M. Immunology of the ageing kidney. Nat Rev Nephrol. 2019;15(10): 625–640.

32 Hsiao YT, Shimizu I, Yoshida Y, Minamino T. Role of circulating molecules in age-related cardiovascular and metabolic disorders. Inflamm Regen. 2022;42(1): 2.

33 Piccoli GB, Cederholm T, Avesani CM, Bakker SJL, Bellizzi V, Cuerda C, et al. Nutritional status and the risk of malnutrition in older adults with chronic kidney disease - implications for low protein intake and nutritional care: A critical review endorsed by ERN-ERA and ESPEN. Clin Nutr. 2023;42(4): 443–457.

34 Yang G, Waheed S, Wang C, Shekh M, Li Z, Wu J. Exosomes and Their Bioengineering Strategies in the Cutaneous Wound Healing and Related Complications: Current Knowledge and Future Perspectives. International Journal of Biological Sciences. 2023;19(5): 1430–1454.

